# Meta-analysis of the SARS-CoV-2 serial interval and the impact of parameter uncertainty on the COVID-19 reproduction number

**DOI:** 10.1101/2020.11.17.20231548

**Authors:** Robert Challen, Ellen Brooks-Pollock, Krasimira Tsaneva-Atanasova, Leon Danon

## Abstract

The serial interval of an infectious disease, commonly interpreted as the time between onset of symptoms in sequentially infected individuals within a chain of transmission, is a key epidemiological quantity involved in estimating the reproduction number. The serial interval is closely related to other key quantities, including the incubation period, the generation interval (the time between sequential infections) and time delays between infection and the observations associated with monitoring an outbreak such as confirmed cases, hospital admissions and deaths. Estimates of these quantities are often based on small data sets from early contact tracing and are subject to considerable uncertainty, which is especially true for early COVID-19 data. In this paper we estimate these key quantities in the context of COVID-19 for the UK, including a meta-analysis of early estimates of the serial interval. We estimate distributions for the serial interval with a mean 5.6 (95% CrI 5.1–6.2) and SD 4.2 (95% CrI 3.9–4.6) days (empirical distribution), the generation interval with a mean 4.8 (95% CrI 4.3–5.41) and SD 1.7 (95% CrI 1.0–2.6) days (fitted gamma distribution), and the incubation period with a mean 5.5 (95% CrI 5.1–5.8) and SD 4.9 (95% CrI 4.5–5.3) days (fitted log normal distribution). We quantify the impact of the uncertainty surrounding the serial interval, generation interval, incubation period and time delays, on the subsequent estimation of the reproduction number, when pragmatic and more formal approaches are taken. These estimates place empirical bounds on the estimates of most relevant model parameters and are expected to contribute to modelling COVID-19 transmission.

## Introduction

Since the end of 2019, the novel strain of coronavirus, SARS-Cov-2 has caused a global pandemic of disease. The speed with which the virus spreads is dependent on biological determinants that enable viral replication within individuals and onward transmission to others. The minimal set of parameters required for understanding the dynamics of any novel infectious disease pathogen include the potential for transmission, the duration of infectiousness (often captured in models as a recovery rate) and the generational interval: the time between two subsequent cases in a chain of infection.

Although much more is understood about these basic parameters than at the beginning of the pandemic, substantial uncertainty remains over the future trajectory of the disease which depends partly on the interventions that are implemented.

The best possible evidence on each parameter determining the dynamics of the epidemic is crucial for choosing the most appropriate interventions. Estimating the generation interval is particularly challenging due to the fundamentally hidden nature of transmission events. In Figure 1 we summarise the timeline of key events for two adjacent infected individuals (infector and infectee) in a chain of transmission.

**Figure 1:**
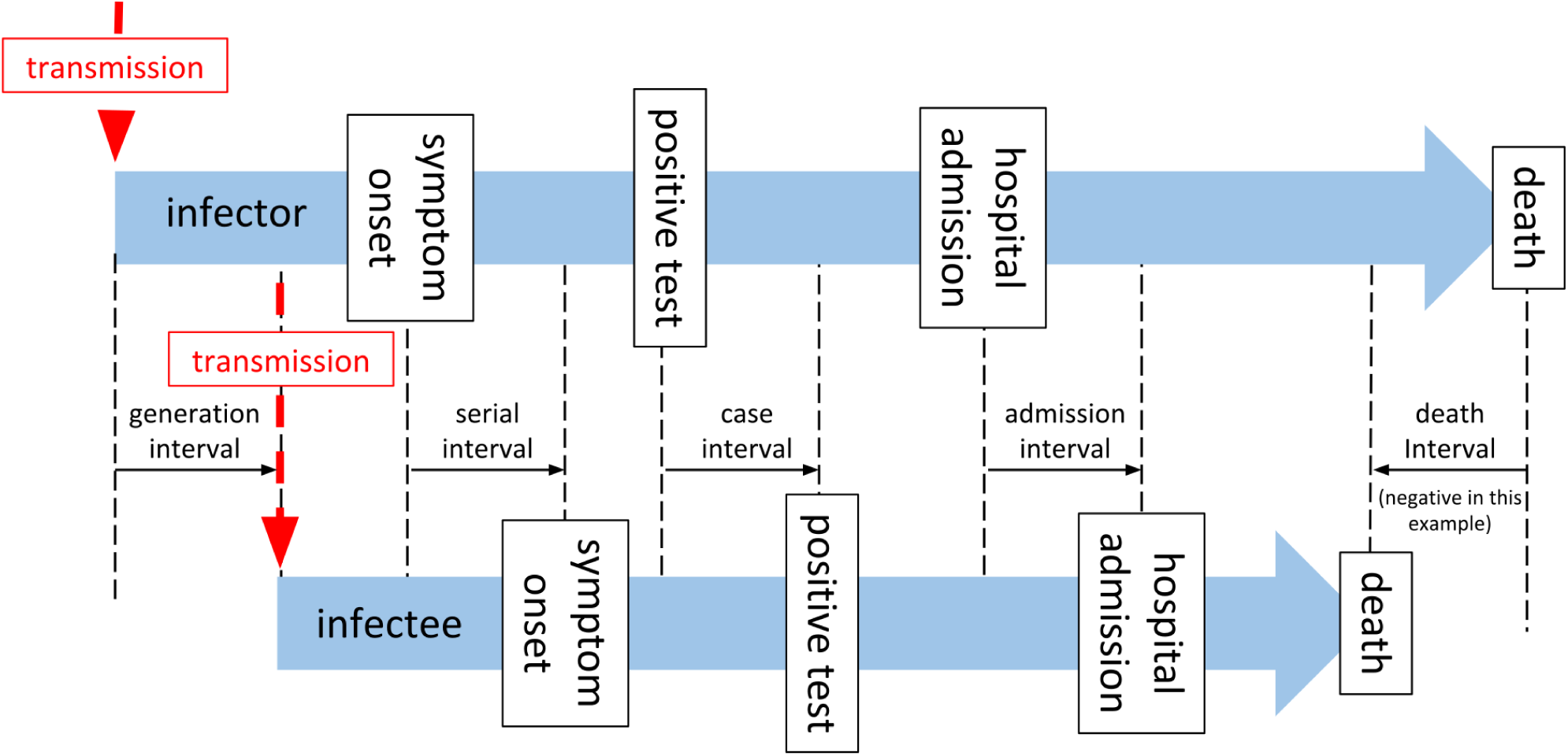
A timeline of events associated with a single infector-infectee pair in a transmission chain.

As described by Svensson et al.^1^ the generation interval is defined as the time between the infection of an infector and infectee, and in practice is not easy to observe, as an infection goes through some latent period, during which it is undetectable^2^, and a pre-symptomatic phase during which an individual may be infectious, and possibly detectable through screening, before the disease manifests with clinical symptoms. The latent period and pre-symptomatic phase are together usually referred to as the incubation period (*T*_*incubation*_). From the onset of symptoms, the diagnosis will be confirmed by some canonical test after some time delay (*T*_*onset→test*_), later the patient may require admission to hospital (*T*_*onset→admission*_), or may die (*T*_*onset→death*_). These subsequent events clearly may not happen in that order, and even diagnosis may occur after death. The time between these key events (onset of symptoms, test confirmation of case, hospital admission, and death) for two sequentially infected people in a chain of transmission are described as serial intervals^1^, although in general usage, and in the rest of this paper, the term “serial interval” is taken to mean the interval between onset of symptoms (*SI*_*onset*_). In Figure 1 and the rest of this paper we use the terms “case interval” (*SI*_*case*_), “admission interval” (*SI*_*admission*_), and “death interval” (*SI*_*death*_) to describe the other intervals. The generation interval is by definition a non-negative quantity, but all the other measures may be negative if the variation of the delay from the event of infection from person to person exceeds the period between infections. This is more likely for death interval than for serial interval, and for diseases with long pre-symptomatic periods, for example HIV^3^.

The reproduction number (*R*_*t*_) is a key measure of the state of the epidemic, and estimates of *R*_*t*_ can become a determinant of public health policy. The “renewal equation” method for estimating *R*_*t*_, depends on a time series of infections, and on the infectivity profile - a measure of the probability that a secondary infection occurred on a specific day after the primary case, given a secondary infection occurred^4,5^. A Bayesian framework is then used to update a prior probabilistic estimate of *R*_*t*_ on any given day with both information gained from the time series of infections in the epidemic to date and the infectivity profile to produce a posterior estimate of *R*_*t*_. In both the original^6^ and revised^5^ implementations of this method, the authors acknowledge the pragmatic use of the serial interval distribution, as a proxy measure for the infectivity profile, and the incidence of symptom onset or case identification as a proxy for the incidence of infection, with the caveat that these introduce a time lag into the estimates of *R*_*t*_. It has been noted by various authors that use of the serial interval as a proxy for infectivity profiles is a pragmatic choice^5,6^ but can introduce a bias into estimates of *R*_*t*_^7,8^.

In the COVID-19 outbreak a limited number estimates of the serial interval are available from studies of travelers from infected areas, and early contact tracing studies (detailed in Table 1). The infectivity profile is comprised of non-negative values by definition. The serial interval, on the other hand, can be measured as negative for several reasons. For example, if the incubation period of the infector is at the short end of the distribution and that of the infectee is at the tail, a negative serial interval would be observed. Negative values have been noted as a feature in at least one estimate of the serial interval of SARS-CoV-2 to date^9^, but cannot be sensibly used in estimates of *R*_*t*_.

**Table 1:**
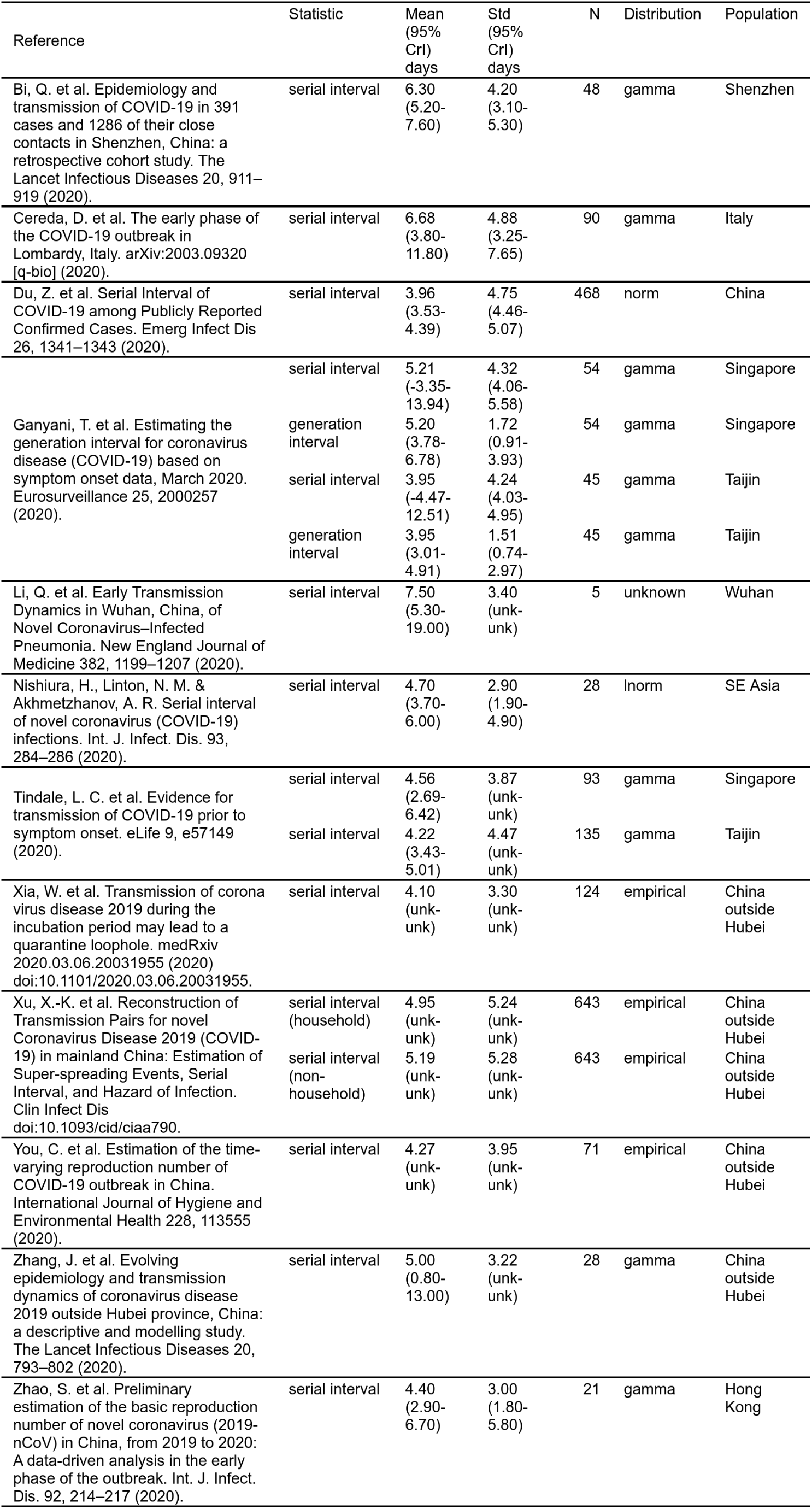
Sources of serial interval estimates from a literature search.

Direct measurement of the serial interval distribution is further complicated by the fact that symptom onset is often not observed due to the scale of the outbreak and that most infection is asymptomatic^10,11^. The data available during an epidemic are a time series of counts of observations, such as confirmed cases (test results confirming diagnosis), hospital admissions, and deaths. As depicted in Figure 1 these events occur after infection, following a period of time. Therefore the observed time series of observed cases is a result of the unobserved time series of infections governed by the serial interval, convolved by the distribution of the time delay from infection to case identification.

Since transmission model-based inference is predicated on infections best practice would be to use the generation interval and infer the unobserved incidence of infection from the observations we have using back propagation or de-convolution^3,8^. A pragmatic alternative^8^ is to simply calculate *R*_*t*_ using a serial interval and un-adjusted case numbers with a simple correction for the time delay between infection and cases by shifting our observed cases in time. To do either of these things we need estimates of the time delays between infection and symptom onset (incubation period), infection and case identification (test), infection and admission, and between infection and death, and their distributions.

The purpose of this paper is to determine the best estimates for the parameters we need to calculate *R*_*t*_ for the UK using either formal or pragmatic approaches. We also wish to understand the degree and nature of bias introduced by using the pragmatic approach using truncated serial interval distributions as approximations for infectivity profile^7^, and case numbers as proxy for infection incidence, compared to the more formal methods of using generation intervals and inferred infection numbers^8^. To do this we investigate estimates for the serial interval, the incubation period, and then time between symptom onset and case identification, admission or death, in the UK. From these we infer the generation interval, and time between infection and case identification, admission and death. With all these parameters available we then compare the impact of using pragmatic and more formal approaches on estimation of *R*_*t*_.

## Methods

### Serial interval estimation

Firstly, we conducted a literature review for studies that describe serial interval estimates using PubMed and the search terms “(SARS-CoV-2 or COVID-19) and ‘Serial interval’” and reviewed the abstracts of relevant original research papers. These were compared to papers reported on the MIDAS Online Portal for COVID-19 Modeling Research^12^, and with existing meta-analyses^13^. From these papers, serial interval mean and standard deviation estimates were extracted, along with information about assumed statistical distributions, and the sample size of the study. A random effects meta-analysis was conducted^14^ on the subset of papers that reported confidence intervals, and which assumed gamma distributions for their estimates of serial interval. This excluded a number of larger studies, and we had concerns over potential violations of the assumptions underpinning the meta-analysis, most notable the assumption of normal distribution of effect size^15–17^, so we also undertook a re-sampling exercise, as described below.

For studies that reported a probability distribution for the serial interval, we randomly selected one hundred probability distributions consistent with those reported in each paper, assuming a normally distributed mean (central limit), and a chi-squared distributed standard deviation with degrees of freedom determined by the sample size of the study. From this family of probability distributions we created random samples based on the original sample size^18,19^. For empirical data reported in the studies we obtained the data where available and used 100 random bootstrap sub-samples with replacement, to a relative size determined by the original sample size of the study. The empirical and distribution based samples were combined into 100 groups of samples, with each group containing representation of each source article with numbers proportional to the size of the original study. A normal, negative binomial and gamma probability distributions were then fitted to these 100 groups using maximum likelihood estimation, implemented in R^20,21^, giving us both parametric probability distribution estimates and 100 empirical probability distributions estimates of the combination of all the source studies (from here on referred to as the “re-sampled serial interval estimate”).

Second, we use data collected under the “First Few Hundred” (FF100) case protocol by Public Health England^22,23^ which provides a limited number of linked cases of proven transmission, mostly within households, and interval censored symptom onset dates. To this data we fitted normal, gamma and negative binomial distributions using the same methodology as above. When fitting gamma and negative binomial distributions we truncated the data at zero, to prevent negative values, and for the gamma distributions we required that the shape parameter had a lower bound of 1, which enforces that the distribution density is zero at time zero.

### Incubation period estimation

The incubation period has been previously estimated by Lauer et al. and Sanche et al.^24,25^ for China in the early phase of the epidemic. The FF100 data contains interval censored exposure data coupled to symptom onset; using this we derived a UK specific estimate to assess if it was consistent. Furthermore, the Open COVID-19 Data Working Group^26,27^ provides a large international data set which includes some travel history and symptom onset data. As the incubation period is a key quantity in the analysis, we reassessed earlier estimates^24,25^ of the incubation period with this larger data set. For both datasets we fitted gamma, weibull, log normal, and negative binomial probability distributions to the intervals between putative exposure and symptom onset, accounting for censoring where present, to estimate the incubation period distribution.

### Generation interval estimation

The generation interval is the fundamental variable for modelling transmission. Under the assumption that it follows a gamma distribution we can infer its parameters using the serial interval (*SI*_*onset*_) distribution, the incubation period distribution, and the constraint that the generation interval is a non-negative quantity. Using the best estimate of the incubation period distribution, we combined random samples from a parameterised probability distribution for the generation interval with two samples from the incubation period to satisfy the following relationship, thus simulating the serial interval:

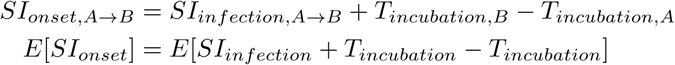

The mean and standard deviation of the simulated serial interval distribution were then compared to the empirical re-sampled serial interval distributions we estimated in an earlier stage. The parameters for the generation interval distribution were then optimized by a recursive linear search on the standard deviation, with the constraints that the mean of the simulated and empirical distributions must be the same^7^, and the standard deviation must be smaller than the mean (ensuring the gamma function scale parameter is larger than one and hence the density is zero at time zero). The minimization function we employed was the absolute difference in inter-quartile ranges of simulated and observed distributions. This process was repeated for 100 different simulated samples that were compared to the 100 different empirical re-sampled serial interval estimates from the previous stage of our analysis to get confidence intervals on our estimates of the generation interval distribution.

### Impact of using the serial interval on estimation of *R*_***t***_

With various estimates of serial interval and generation interval we wished to understand the qualitative impact this variation might have on our estimates of *R*_*t*_. To investigate this we used the forward equation approach implemented in the R library EpiEstim^4,6,28^. We estimate values of *R*_*t*_ for 4 time points in the first wave of the COVID-19 pandemic in England representative of the ascending phase, the peak, the early descending phase and the late descending phase. We used data retrieved from the Public Health England API^29,30^. For this analysis we assume the infectivity profile can be represented using a parameterised gamma distribution, and estimate *R*_*t*_ for a wide range of combinations of mean and standard deviation, using a fixed calculation window of 7 days, at each of our four time points. The resulting relationship between *R*_*t*_, mean infectivity profile, and standard deviation of infectivity profile were compared visually.

### Time delays from infection to case identification, admission, and death

Estimation of the time interval between the onset of symptoms and the observations of positive test result, hospital admission, and death was performed (*T*_*onset→test*_, *T*_*onset→admission*_, and *T*_*onset→death*_) using the CHESS data set^31^. The CHESS data set is hospital based, and was initially limited to intensive care admissions, but a subset of hospitals have reported all admissions, and this is what we focused on. Within the CHESS data set there are a set of patients who have symptom onset dates recorded, dates that a specimen was taken that subsequently was tested positive, hospital admission date, and date of death, if the patient died. The time delays from symptom onset to the different observations were calculated and fitted to probability distributions using the R library fitdistrplus^21^ as described above.

The time delay from infection to observation was obtained by combining our estimate of the incubation period distribution from the Open COVID-19 Data Working Group data set^26,27^ with onset to observation delays from the CHESS data set^31^ using the following relationship.

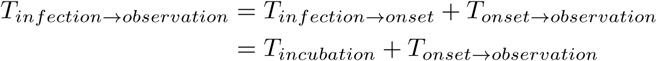

We combined the incubation period and onset to observation distributions using a random sampling approach, assuming independence of the two variables. These random samples were then estimated as parameterised statistical distributions in the same manner as above, with the constraint that all the time delays from infection to observation are non-negative quantities, and their probability is zero at time zero.

### Impact of deconvolution

In the final piece of our analysis we sought to qualitatively compare estimates of *R*_*t*_ based on data for England from the Public Health England API^29,30^, in each of the following two scenarios.

Firstly, we based *R*_*t*_ estimates on observational data (cases, admissions, deaths) as a proxy for infection events, and used truncated serial interval distribution as a proxy for infectivity profile. A simple-but-incorrect adjustment to the dates of these *R*_*t*_ estimates was made to align the estimate to date of infection rather than date of observation.

Secondly, using the time delay distributions from the previous stage, we used de-convolution to infer a set of time series of infections from the same observational data and used our estimate of the generation interval as a proxy for the infectivity profile. This second approach has been recommended by Gostic et al^8^. To do this we applied a non parametric back projection algorithm from the surveillance R package^32^, based on work by Becker et al.^3^ and Yip et al.^33^, to infer three putative infection time series from observed cases, admissions or deaths. The inferred time series were then used to estimate *R*_*t*_ through EpiEstim using the parametric gamma distributed estimate of the generation interval from above. In applying the de-convolution we discovered it requires a full time series beginning with zero cases for sensible results and this required we impute the early part of the hospital admission time series, which we did by assuming an early constant exponential growth phase.

The resulting *R*_*t*_ time series were compared qualitatively.

## Results

### Serial interval estimation

Our PubMed search retrieved 62 search hits of which 12 were original research articles containing estimates of serial intervals^9,34–43^. The mean and standard deviation of parameterised distributions were extracted and are presented in Table 1. The estimates of the mean range from 3.95 days to 7.5 days. The majority of studies provided their results as gamma distributions defined by mean and standard deviation. Some studies, particularly Xu et al.^9^ noted that the serial interval was not infrequently negative.

The random effects meta-analysis on the subset of studies which reported gamma distributions, resulted in an overall estimate of the serial interval that follows a gamma distribution, with a mean plus 95% credible interval of 4.97 days (3.85; 6.07), and a standard deviation of 4.23 days (3.84; 4.61) (for more details see Supplemental figure 1).

In Figure 2, panel A we present the results of the re-sampled serial interval estimate. The histogram shows the empirical distribution of the combination of all the studies, reinforcing the finding of a substantial proportion of the serial interval being negative. For the gamma and negative binomial distribution fit the data is truncated at zero, and the full data used for the normal distribution, resulting with mean values of 5.59 (gamma), 5.53 (negative binomial) or 4.88 (normal) days. Full detail of the parameterisation of this is available in Supplemental table 1.

**Figure 2:**
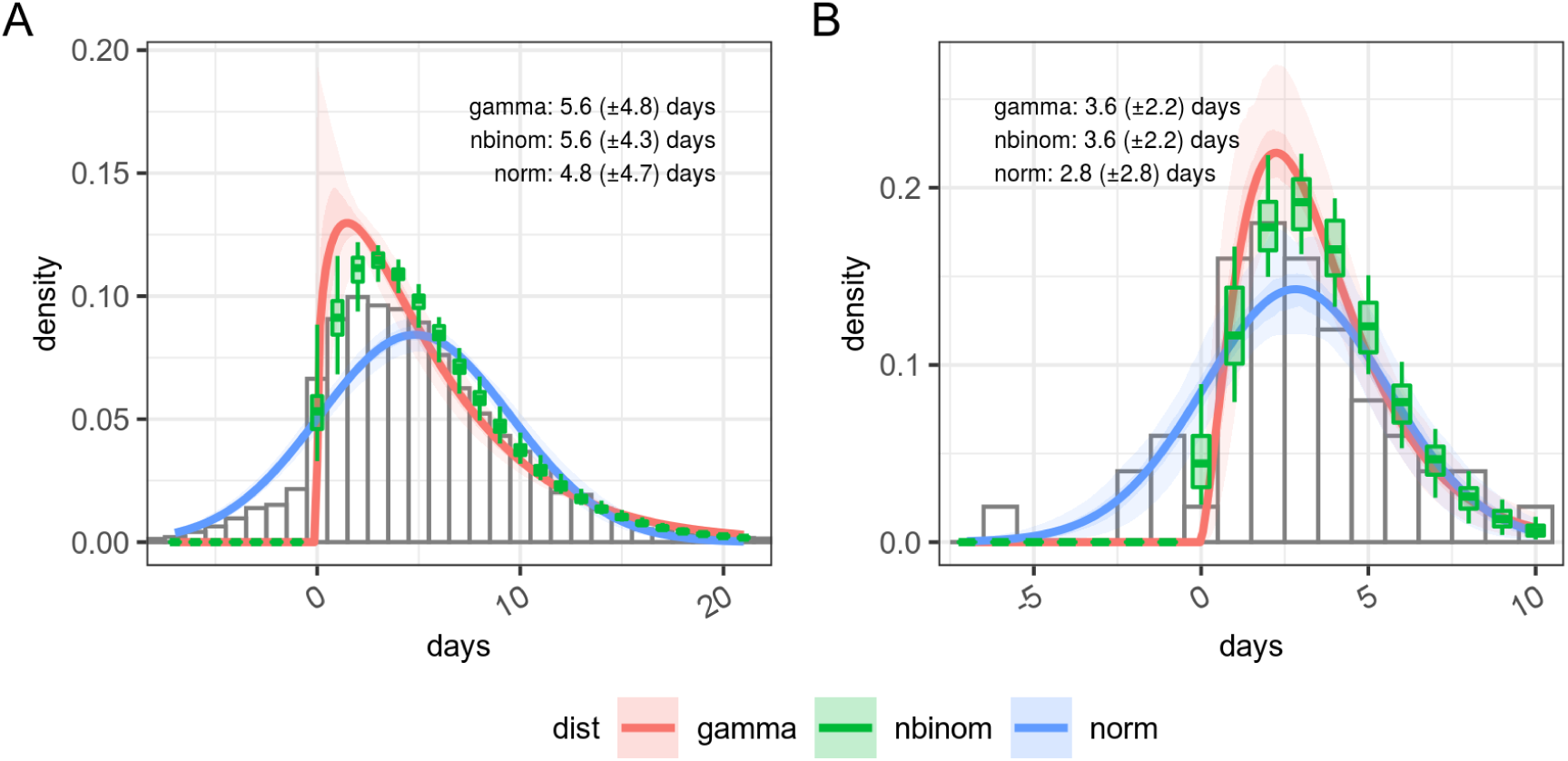
Panel A - Days between infected infectee disease onset based on a resampling of published estimates from the literature and Panel B - Estimates of serial interval from FF100 data. The histogram in panel A shows the combined density of all sets of samples within the original research.

In Figure 2 panel B we present the distribution of the 50 linked cases in the FF100 data set for which onset dates are available for both infector and infectee. As with the resampled data the parameterised versions of this are based on truncated data for negative binomial and gamma distributions, and hence shows a poorer fit against the whole distribution (full detail of the parameterisation of this is available in Supplemental table 2). Supporting the observations of Xu et al.^9^, the FF100 data shows evidence of negative serial intervals. The mean of the serial interval from FF100 data was 3.54 days when parameterised with a gamma distribution and data truncated to exclude negative serial intervals, and 2.09 days when a normal distribution used, with no truncation. This is on the lower end of the values reported in the literature.

**Table 2:** Goodness of fit statistics for incubation period distributions reconstructed from Open COVID-19 Data Working Group and from FF100 data.

| Source | N | AIC | BIC | Log-likelihood | Distribution |
| --- | --- | --- | --- | --- | --- |
| FF100 | 32 | 67.3 | 70.3 | -31.7 | negative binomial |
|  |  | 70.6 | 73.5 | -33.3 | weibull |
|  |  | 70.6 | 73.5 | -33.3 | gamma |
|  |  | 72.3 | 75.3 | -34.2 | log-normal |
| Open COVID-19<br>Data Working<br>Group | 1071 | 5.34e+03 | 5.35e+03 | -2.67e+03 | log-normal |
|  |  | 5.51e+03 | 5.52e+03 | -2.75e+03 | gamma |
|  |  | 5.55e+03 | 5.56e+03 | -2.77e+03 | weibull |
|  |  | 5.66e+03 | 5.67e+03 | -2.83e+03 | negative binomial |

As noted in the methods, the re-sampling process allows us to estimate the serial interval as an empirical distribution. Within EpiEstim, our chosen framework for estimating *R*_*t*_ however the use of negative serial intervals are not supported as a proxy for the infectivity profile. Pragmatically we therefore decided to truncate the empirical distribution at zero for use in EpiEstim. Our adopted estimate therefore has a mode of 4.8 days, in line with the normal distribution parameterisation but is a truncated empirical distribution, with a mean plus 95% credible interval of 5.61 days (5.15; 6.22), and a standard deviation of 4.23 days (3.95; 4.64), which is more in line with the gamma distribution parameterisation.

### Incubation period estimation

Figure 3 and Table 2 show the results of estimating a parametric probability distribution to data from FF100 and data from the Open COVID-19 Data Working Group. Histograms of the data are not shown as it is interval censored, which is not straightforward to represent graphically. There are only a small number of records from the FF100 data which suggest the mean of the incubation period is between 1.8 and 2 days. The data from the Open COVID-19 Data Working Group suggests the incubation period is longer with a mean around 5.5 days depending on the distribution chosen, and this agrees better with other estimates in the literature^13,24,25^. The best fit to the Open COVID-19 data is obtained with a log normal distribution as seen in Table 2 (Full details of the fitting parameters are in Supplemental table 3).

**Table 3:** Estimated time delays between infection and various observations over the course of an infection, based on the combination of incubation period and symptom onset to observation delay.

| Observation | Mean delay (days) | SD (days) | 95% quantiles (days) |
| --- | --- | --- | --- |
| onset | 5.42 | 5.89 | 0.64; 20.86 |
| test | 7.80 | 7.23 | 0.74; 26.05 |
| admission | 9.23 | 10.64 | 0.80; 38.32 |
| death | 17.29 | 12.40 | 3.66; 49.09 |

**Figure 3:**
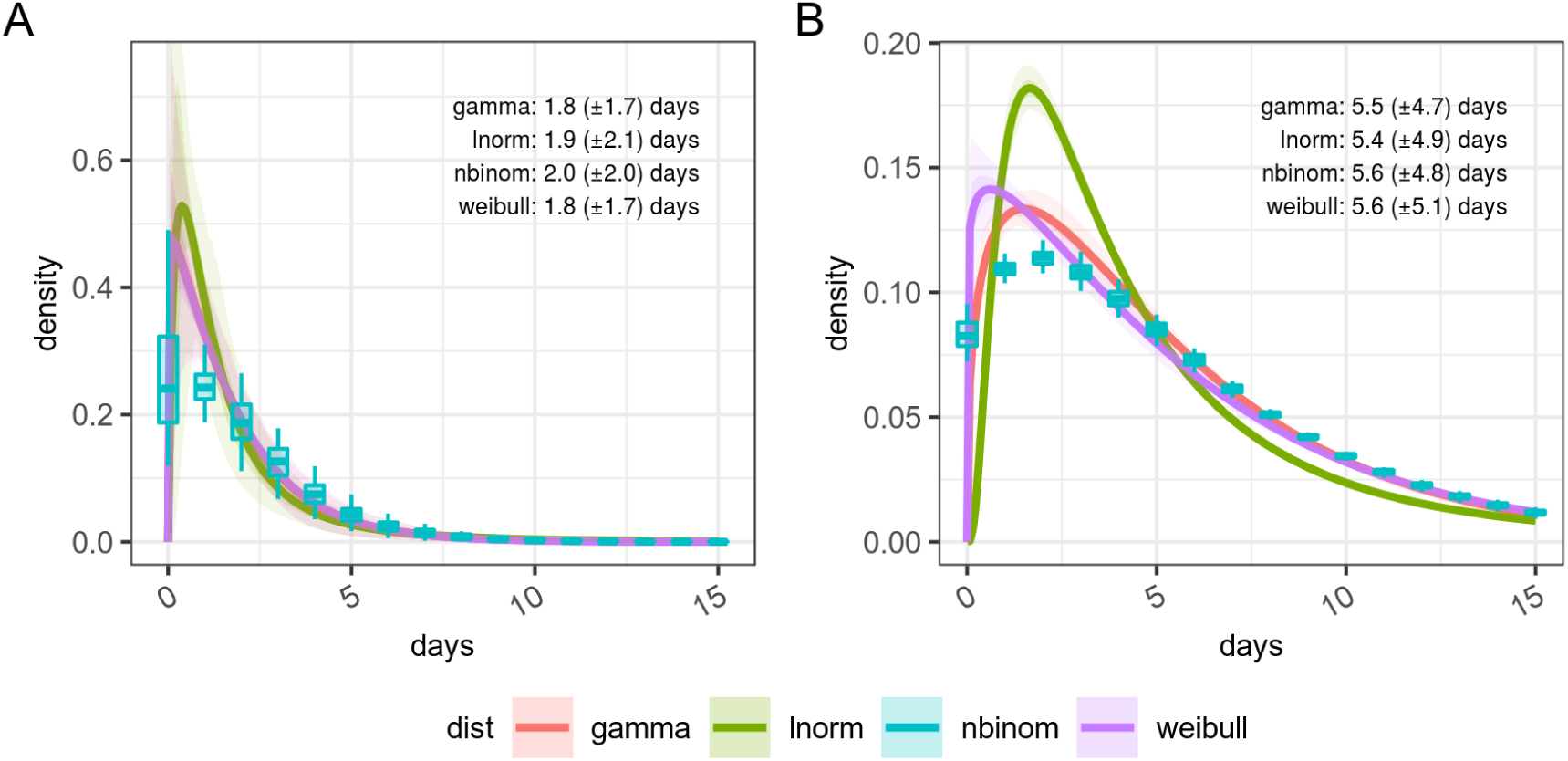
Incubation period distributions reconstructed from Open COVID-19 Data Working Group and from FF100 data. Histogram data is not shown as the interval censored data cannot be plotted in this form.

### Generation interval estimation

The generation interval is then inferred from the incubation period and empirical serial interval distribution prior to truncation. Our best estimate for this is a gamma distribution, with a mean plus 95% credible interval of 4.83 days (4.31; 5.40), and a standard deviation of 1.73 days (0.98; 2.55), as shown in Figure 4. The mean of 4.8 days is identical to that of the empirical serial interval distribution prior to truncation in panel B, Figure 2 as a result of the constraints imposed during fitting. The standard deviation of our generation interval estimate is 1.72. This is within the confidence limits of estimates from the literature from both China (0.74 - 2.97) and Singapore (0.91 - 3.93)^44^.

**Figure 4:**
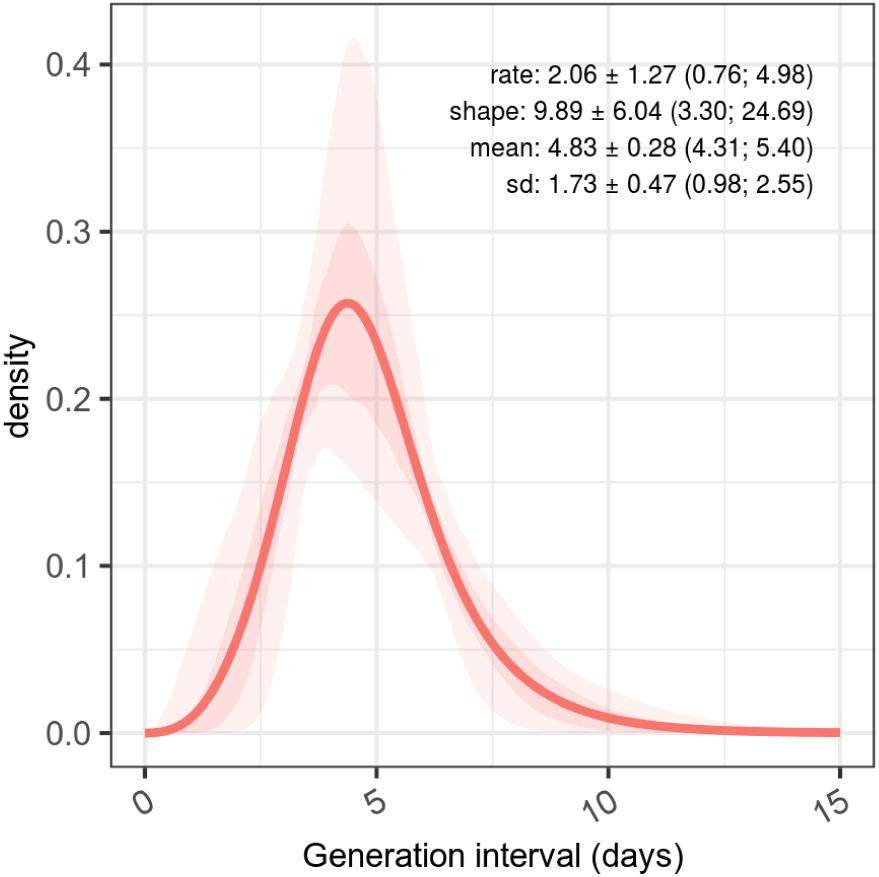
Estimated generation interval distributions, from resampled serial intervals as predictor, and estimated serial intervals from incubation period combined with samples from a generation interval assumed as a gamma distributed quantity.

### Impact of using the serial interval on estimation of *R*_***t***_

With our 3 estimates of the serial interval and 1 generation interval and observed COVID-19 case counts, we investigate the impact on the estimates of *R*_*t*_, of using these estimates as a proxy for the infectivity profile. This uses data at 4 time points on an epidemic curve from the first wave of the COVID-19 outbreak in England, shown in Figure 5, which are 19th March, 12th April, 12th May and 23rd June, corresponding to the ascending, peak, early descending and late descending phases respectively.

**Figure 5:**
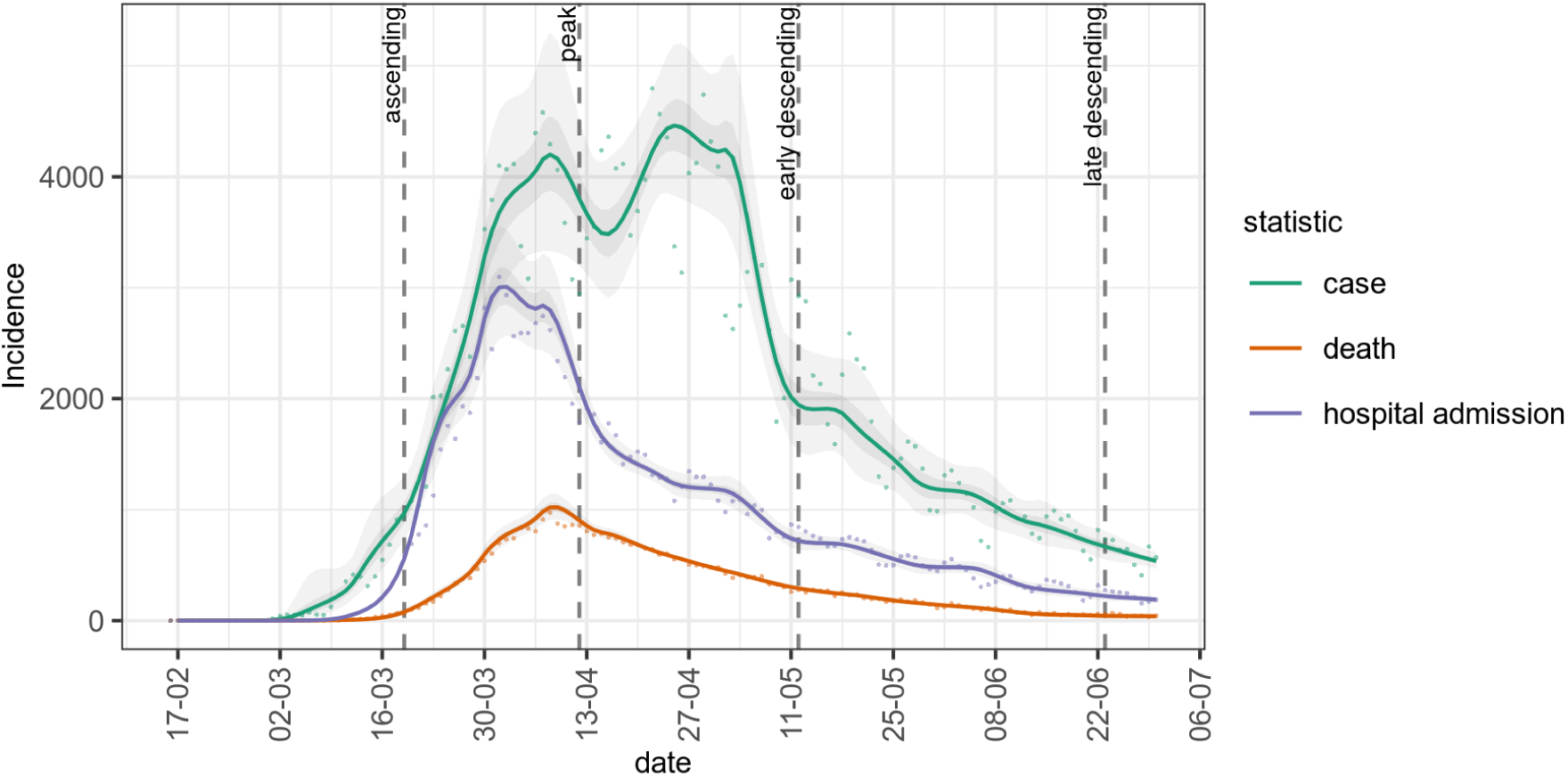
Epidemic curve for cases, deaths and hospital admissions are used for analysis in this paper. Dashed vertical lines show dates at which we conduct our analysis, chosen to represent the ascending, peak, early and late descending phases of cases during the first wave in the UK.

At each of these 4 time points Figure 6 shows the estimated *R*_*t*_ under the range of different assumptions about the mean and standard deviation of the infectivity profile, modeled as a gamma distribution. In the top left, bottom left and bottom right panels the effect of increasing the mean of the infectivity profile is to push the resulting estimate of *R*_*t*_ away from the critical value of 1 at which the epidemic is growing. In the top right panel, at the peak, the mean of the infectivity profile has a less clear-cut effect. The impact of changes to standard deviation is likewise varied. In the top left, bottom left and bottom right panels during ascending and descending phases there is relatively little impact of changing the standard deviation of the infectivity profile on the estimates of *R*_*t*_, and any small changes that do occur depend on the shape of the preceding epidemic curve. At the peak however, in the top right panel, the wider the standard deviation the more historical information influences the estimation of *R*_*t*_ and this acts to delay the estimated transition from positive to negative growth. The overall result of this is that estimates of the infectivity profile with a high standard deviation will predict *R*_*t*_ crossing 1 later than estimates based on an infectivity profile with a low standard deviation, but the point of crossing 1 is relatively insensitive to the value of the mean of the infectivity profile.

**Figure 6:**
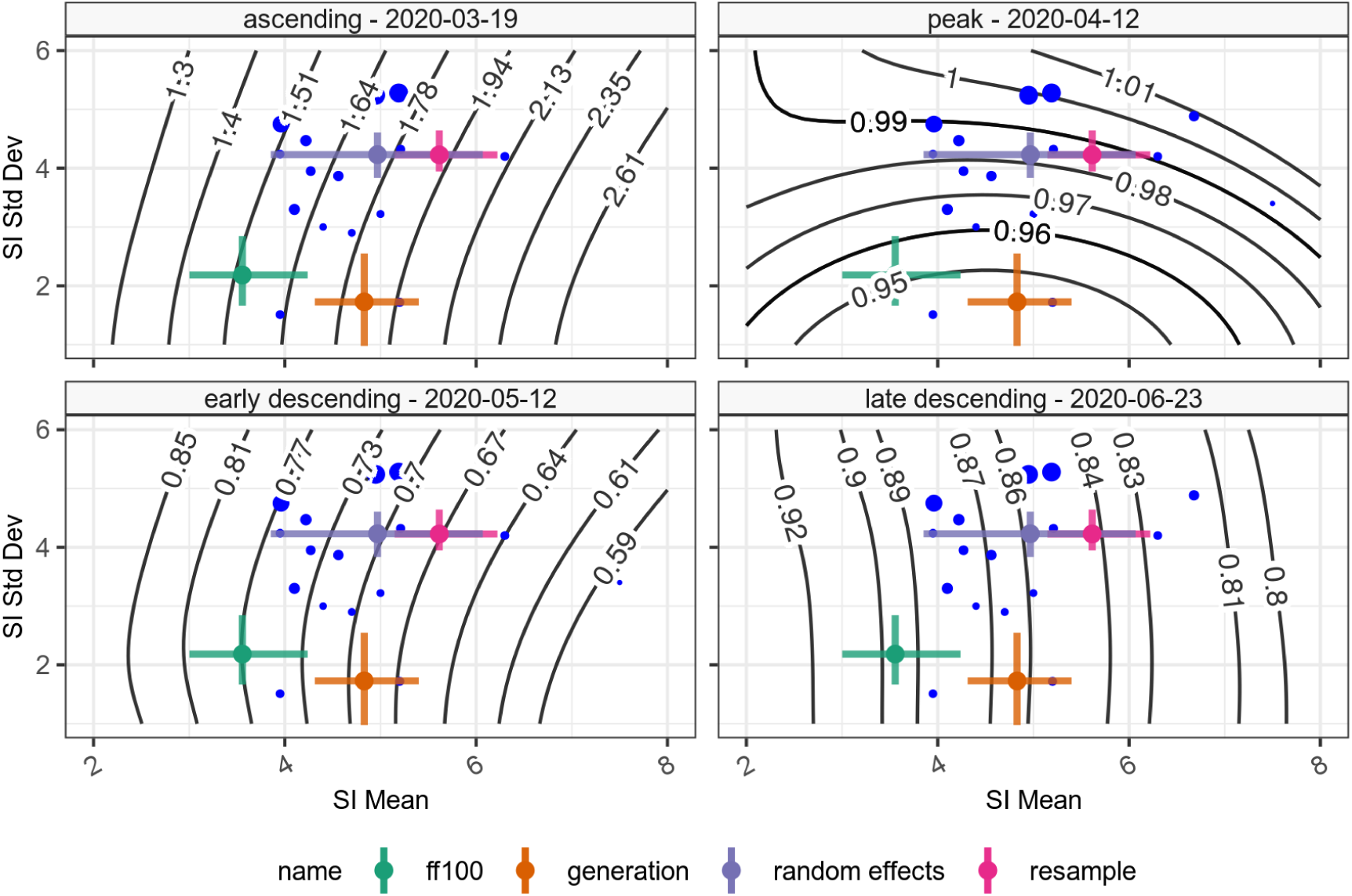
Time varying reproduction number given various assumptions on the serial interval mean and standard deviation. The blue points show the central estimate of serial intervals from the literature, whereas the coloured error bars show the mean and standard deviation of the 3 serial interval (green, violet, magenta) and 1 generation interval (orange) estimates presented in this paper. Contours show the *R*_*t*_ estimate for that combination of mean and standard deviation serial interval. The four panels represent the 4 different time points investigated.

When we consider using the various estimates of the serial interval or generation interval, as a proxy for the infectivity profile, on the resulting estimates of *R*_*t*_ we can see from the coloured crosses on Figure 6 representing the different estimates of serial or generation interval, that in the situations of dynamic change such as the ascending phase the variability may have quite a large impact on subsequent estimates of *R*_*t*_ but at other times the impact is much smaller [ascending - 21% variation (*R*_*t*_: 1.52 to 1.88); peak - 4% variation (*R*_*t*_: 0.94 to 0.98); early descending - 13% variation (*R*_*t*_: 0.67 to 0.77); late descending - 6% variation (*R*_*t*_: 0.84 to 0.90)].

### Time delays from infection to case identification, admission, and death

In Figure 7 we show probability distributions fitted to data from the CHESS data set which define times from symptom onset (Panel A) to case identification (*T*_*onset→case*_), admission (*T*_*onset→admission*_), or death (*T*_*onset→death*_). Symptom onset to test (case identification) can be a negative quantity if a swab is taken during disease screening and the patient is pre-symptomatic. In this data the time point that defines the time of test is the date when the specimen is taken, which will subsequently be tested positive for SARS-CoV-2, so does not include sample processing delays. However in this hospital based data source of admitted patients the onset data was collected retrospectively. We also note peaks at 1 day, 1 week, 2 weeks, and so on which suggests approximation on data entry, and there may well be biases in the data collection.

**Figure 7:**
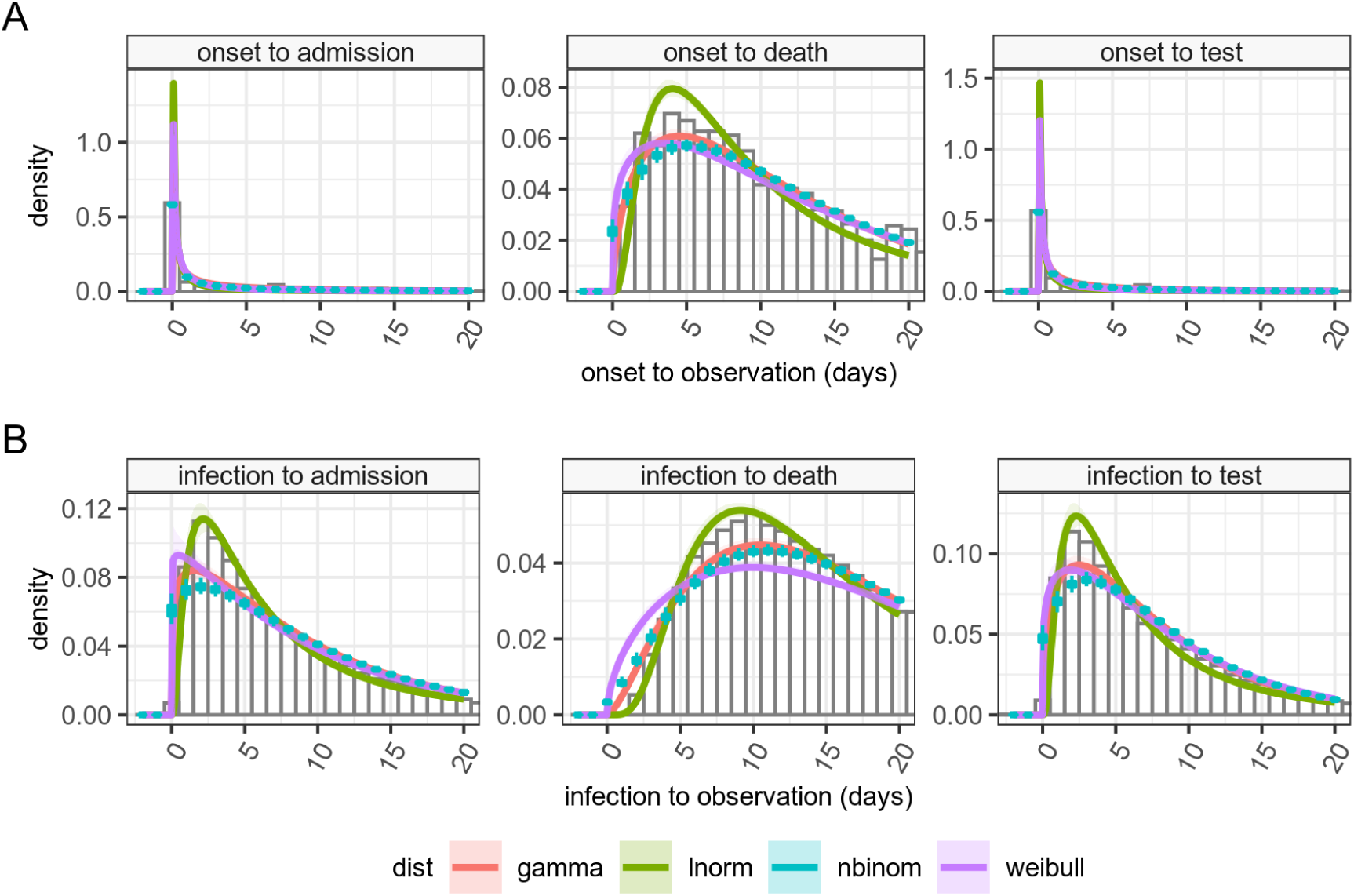
Panel A: Time delay distributions from symptom onset to test (diagnosis or case identification), admission or death, estimated from CHESS data set, plus in Panel B estimated delays from infection to observation, and can be negative in certain cases. based on incubation period and observation delay. These can be used for deconvolution.

It is more obvious from the clinical course of COVID-19 that admission should occur after disease onset. In the data, a large number of cases are reported to have symptom onset on day of admission. This is potentially a reporting artifact as in the absence of certain knowledge about onset, it is possible that the day of admission may have been captured instead, and we again see the peaks at 1 week, 2 weeks, and so on, suggesting approximations in data entry.

The time between onset of symptoms and death can also be assumed as a positive quantity given this is based on an in hospital cohort. This distribution shows a large tail, and some patients in that tail were noted to be admitted many months before the appearance of COVID-19. These patients likely represent hospital acquired cases in chronically unwell patients. The extreme outlying values (with delay from admission to death greater than 100 days, or with an admission before 1st Jan 2020) were removed as they prevented sensible estimation of the rest of the distribution.

By combining the incubation period distribution in panel B in Figure 3 with the time delay distributions in panel A of Figure 7, we can obtain probability distributions from infection to observation, and these are shown in panel B for the 3 observations of test (case identification), admission and death. These distributions provide us with a means of estimating a time series of infection from observed case counts, admissions and deaths. As above full details of their parameterisations are available in Supplemental table 4. The mean time from infection to the various time points described in the timeline in Figure 1 are presented in Table 3. The infection to onset is the incubation period, with a mean of 5.5 days. On average approximately 8 days pass from infection to diagnosis, a subsequent 1 days until admission, and a further 7-8 days until death, however it also shows considerable variation in these delays, exemplified by the 95% quantiles for the time from infection to death estimated as ranging from 3.6 days to nearly 50 days.

### Estimation of *R*_*t*_

With the estimates of delay from infection to observation we are able to use a non parametric back propagation as described in the methods to estimate a time series of infections as recommended by Gostic et al.^8^ when using EpiEstim. The results of the back propagation is shown in Figure 8, panel A which depicts resulting point and smoothed infection curves associated with observation curves from England. The back projection results in a sharper and narrower epidemic curve than the observation it is derived from, and indicates additional structures (such as more pronounced fluctuations) which are not obvious from the underlying observations. Estimates of *R*_*t*_ are shown in panel B based on de-convolved time series plus generation interval, versus raw observation counts, re-sampled serial interval estimates, with time adjustment on the resulting *R*_*t*_ estimates to align *R*_*t*_ estimate date to putative date of infection. We have not calculated confidence intervals for the estimates of *R*_*t*_. The estimates of *R*_*t*_ differ from their mean (using symmetric mean absolute percentage error, sMAPE) by between 6% to 9% [case: sMAPE 6.97% (IQR 4.06%; 15.79%); death: sMAPE 11.02% (IQR 5.66%; 29.31%); hospital admission: sMAPE 12.02% (IQR 5.96%; 29.92%)].

**Figure 8:**
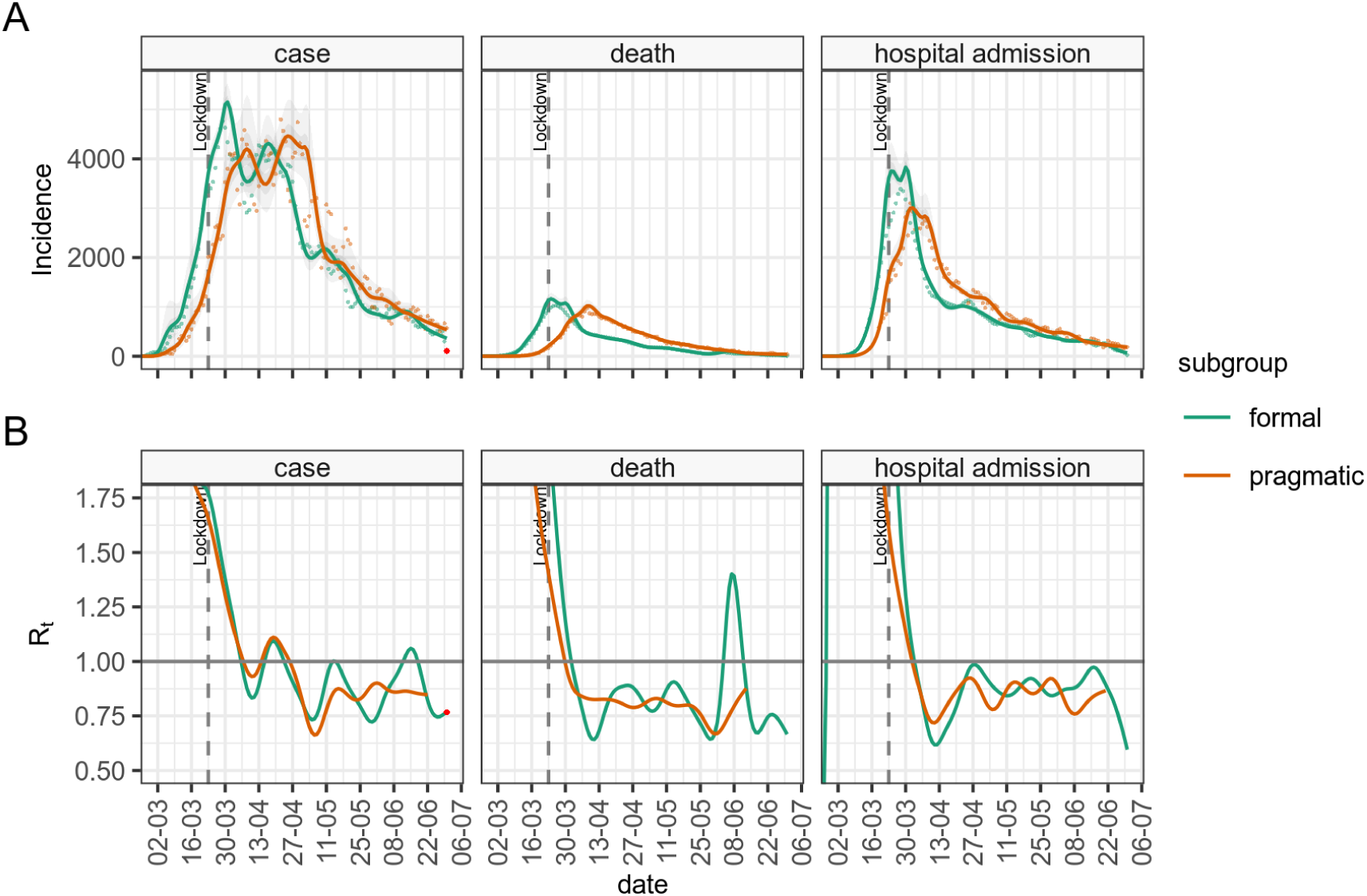
Panel A: The epidemic incidence curves in England for different observations (orange - formal) and inferred estimates of infection rates (green - pragmatic) based on a deconvolution of the time delay distributions. Panel B, the resulting *R*_*t*_ values calculated either using infection rate estimates and generation interval (formal subgroup) or unadjusted incidence of observation, and serial interval (pragmatic). The *R*_*t*_ estimated direct from observed incidence curves (pragmatic) have their dates adjusted by the mean delay estimate.

## Discussion

Our estimate of the serial interval from the FF100 UK data was found to be short, compared to international estimates. The FF100 data was collected in the early stage of the epidemic and is based mainly on household contacts of international travelers which may impart bias. As the participants in the study were put into self-isolation upon discovery observations the contact period tends to be shortened, leading to shorter serial interval estimates, and because of this we do not believe the estimate from the FF100 study to be specific to the UK but rather that the data set is not broadly representative of the UK population.

In contrast, our literature search allowed us to estimate the serial interval from a much larger pooled sample drawn from multiple studies. A meta-analysis and a re-sampling study produced comparable results (meta-analysis: mean 5.0 (95% CrI 3.9–6.1) and SD 4.2 (95% CrI 3.8–4.6) days and re-sampling: mean 5.6 (95% CrI 5.1–6.2) and SD 4.2 (95% CrI 3.9–4.6) days), but our re-sampling study more clearly showed the potential for the serial interval to be negative, due to the relatively long and variable incubation period of SARS-CoV-2. The negative values of the serial interval are theoretically problematic for their use as a proxy for the infectivity profile of SARS-CoV-2 in transmission modelling, which requires the infectee is infected after the infector. Pragmatically, as a way around this, we may truncate the re-sampled serial interval at zero, and use this as the infectivity profile, but as seen here this truncation increases the mean of the resulting serial interval distribution from 4.8 to 5.6 days.

An alternative to this is to use the generation interval as a proxy for the infectivity profile, but there are limited estimates of this quantity available in the literature^44^. Derivation of the generation interval from the serial interval is possible using knowledge of the incubation period. We again looked at the FF100 data for estimates of incubation period and again found that the UK data suggests a value which is shorter that international estimates, for the same reasons as mentioned above^24,40,45–47^. We cross referenced this with a second estimate derived from the Open COVID-19 Data Working Group data set^26,27^, which is based on a large international data set of people who tested positive for SAR-CoV-2 after travelling from areas with outbreaks. The resulting estimates of the mean incubation period of 5.5 days (log normally distributed) is much closer to previous estimates, and we expect to be less influenced by right censoring. Again, we regard the short incubation period calculated from the FF100 data set to be a feature of the data set rather than a UK specific finding. In fact the estimate based on the Open COVID-19 Data may in itself be an under-estimate as the majority of travel histories only include the return date of the visit in question and not the start date.

With incubation period and serial interval estimates, we derived an estimate for generation interval, assuming a gamma distribution. This was comparable to previous estimates in that although it is based on a different serial interval, and hence has a different mean, the standard deviation of our estimate is in accordance with that estimated by Ganyani et al.^44^ Although the confidence intervals for the mean and standard deviation of the generation interval are comparable, the variation in the shape and rate parameters of the underlying gamma distribution are quite large. Care should be taken when generating bootstrap samples from the generation interval when it is specified as an uncertain gamma distribution parameterised with mean and standard deviation, as it is possible that some combinations of parameters produce unrealistic distributions, particularly when the standard deviation is small and the mean is large, which could in theory result in posterior estimates for *R*_*t*_ being largely determined by the reciprocal of just a small number of observations.

Using estimates of the serial interval distribution and generation interval distribution as a proxy for infectivity profile, we investigated the resulting variation in the estimation of *R*_*t*_ using incident cases in England and the forward equation approach. We found the bias between the smallest and largest of our estimates to be as high as 20-25% of the central estimate of *R*_*t*_ when *R*_*t*_ was high, but somewhat smaller when *R*_*t*_ values were 1 or lower. Distributions with lower values of the mean tended to result in estimates of *R*_*t*_ that were closer to one. This suggests that biases introduced by the use of different serial interval distributions should not influence the answer to the key question, “is *R*_*t*_ greater or less than 1?” which defines whether the epidemic is expanding or contracting in size. It is also to be expected that the nature of change of *R*_*t*_ over time is not affected by this bias, so an increasing value of *R*_*t*_ will be increasing regardless of the infectivity profile that is used to estimate it.

Estimating *R*_*t*_ is based on knowledge of the incidence of infections. This is not a quantity that is readily observed in the SARS-CoV-2 epidemic, and pragmatically use of observations including symptom onset, case identification, admission and death are expected to be used as proxy measures for infection^6,28^. However, as pointed out elsewhere^8^ the variable time between infection and such observations causes the signal to be both delayed and blurred. By combining our estimates of incubation period with data from hospital admissions in the CHESS data set we were able to make estimates of the distribution of the delay from infection to observation for the UK. There are several caveats to this part of our analysis that must be kept in mind. The CHESS data set relies on a retrospective report of onset of symptoms, and this field is not recorded for all patients. We select only those patients who have reported an onset date and it may be the case that these patients represent a subgroup of patients whose symptom onset is significantly different from the average patient. The data collection around these dates was noted above to show patterns suggestive of rounding or approximation and this could also introduce some bias. The delay distributions are also unlikely to remain fixed during the outbreak, as we would hope to see the time from infection to case identification shorten during the epidemic, and the time from infection to death lengthen as treatment improves, and as the cohort of susceptible individuals changes. Our confidence in these distributions is therefore somewhat low, although we note acceptable agreement between our estimates produced using a mixture of international and UK data, and previously published estimates from different countries, most notably China^47,49–51^ which cite an onset to admission of 2.7-5.9 days^39,47,52^ and onset to death delay of 16.1-17.8 days^47,52^. Given our estimate of the incubation period is log normally distributed, it is unsurprising that the combination of incubation period and delay from onset to observation is also best described by log normal distributions (Figure 7, panel B), and with these we can apply non parametric back propagation to infer a time series of putative infections from the delayed observations we have available.

In Figure 8 we bring all the different parts of the analysis together and compare the two approaches of formal estimation of *R*_*t*_ using the generation interval and back propagation of case, admission or death counts to putative infection, versus an pragmatic estimation using the truncated re-sampled serial interval, and direct use of observation numbers, combined with the simple-but-incorrect adjustment to the resulting *R*_*t*_ time series, shifting the date backwards by the mean of the delay distribution^8^.

In comparing formal versus pragmatic methods, given the number of moving parts in this comparison it is encouraging to see the level of agreement between the *R*_*t*_ estimates from the two methods (Figure 8 panel B) in the early phase of the epidemic, and also to see that there is some similar structure of the *R*_*t*_ time series in the later parts. This is particularly the case for estimates based on cases and admissions, but less so for deaths where the de-convolution time series shows additional features that are not obvious from the data. We have no gold standard in comparing the *R*_*t*_ estimates for England, so we are limited in what we can conclude, but we do observe that *R*_*t*_ estimates based on our attempt at de-convolution have more variability than ones from un-adjusted observations, and de-convolved estimates have additional features not present in the un-adjusted estimates. The de-convolved estimates of infection are also noted to run to the end of the time series, which is somewhat surprising as estimates of infection rates at the end of the time series should depend on data which has not yet been observed. This is a feature of the back propagation algorithm which needs to be used with caution as the resulting estimates of *R*_*t*_ based on de-convolution for the latter part of the time series appear inconsistent with each other.

## Limitations

We did not fully quantify uncertainty in our analysis and estimation of *R*_*t*_. The informal approach to estimating *R*_*t*_ has uncertainty arising from the serial interval distribution, and stochastic noise in the value of the observation in question. The formal approach involves uncertainty in the initial estimation of the serial interval, uncertainty in the incubation period, resulting in uncertainty in the generation interval, the back propagation itself is a source of uncertainty and involves uncertain time delay distributions which are in turn based on uncertain incubation period, and finally the stochastic noise in the observation under consideration. Accurately tracking the uncertainty of all these components into a final estimate remains a challenge.

Our estimates are based on the best available information at the present stage of the epidemic. However the serial interval is not a fixed quantity and may be affected by behavioral changes such as case isolation, or social distancing. The assumption it is constant is questionable although we have very little hard evidence about how it may vary over time. Similarly time distributions from infection to case identification, admission and death are expected to be highly variable over the course of an outbreak. This has implications for their use in de-convolution, as changing time distributions will have a significant effect on the shape of inferred infection incidence curves, and the complexity of the de-convolution.

There are implicit selection biases in all the data sources we use. A large proportion of SARS-CoV-2 cases are asymptomatic^11,53,54^. It is highly likely that these people participate in transmission chains, and they may do so with a very different infectivity profile to those that are symptomatic, however we have no information about these people in the data sets, and hence all the estimates presented here could be quite different, when asymptomatic cases are taken into consideration.

Our approach in estimating key parameters has been to combine different data sets, which come from different international sources, and which have potentially different biases. In combining data sets we assume that time delays are independent of one another, and can be combined randomly, as we have no other evidence to the contrary. This assumption is questionable, as physiologically we can imagine that patients with a long incubation period, for example, may well have a longer period from symptom onset to admission, for example. This could have unpredictable effects on our estimates of time delays but the most likely is that our estimated variance is too small as a result.

## Conclusions

We argue that our estimates for the statistical distributions of these key parameters or serial interval, incubation period, generation interval, for the UK, along with their uncertainty represent the best available estimates for the UK, given the current state of knowledge.

As there is a wide range of candidate values for these quantities, we have assessed the bias that the variation in choice of parameters introduces to *R*_*t*_ estimation, when using the forward equation method. Whilst these introduce significant variation in the worst case scenario, we find that even large differences in infectivity profile can have only small impacts on estimates of *R*_*t*_ when *R*_*t*_ is close to 1. Larger values for the mean of the infectivity profile appear to result in *R*_*t*_ estimates that are further away from 1. This is a relatively reassuring finding in that the answer to the key question “is the epidemic under control?” is insensitive to the mean of the infectivity profile.

Using more formal methods for estimating *R*_*t*_ by back propagation inference of infection rate and estimates of the generation interval, produces *R*_*t*_ estimates that are more variable when compared to the more pragmatic direct use of case counts as a proxy for infection. Both methods agree on when the epidemic crossed the *R*_*t*_ threshold of 1. However there is considerable uncertainty in the quantities needed to perform the back propagation, and we are not able to ensure that all this uncertainty could be faithfully quantified in our resulting *R*_*t*_ estimates. We did not set out to assess whether one method is better than another, and this would be a natural extension to this work, however we note that new methods for combining back propagation with estimation of *R*_*t*_ are under active development^55,56^ and may very well address such further questions.

## Data Availability

All code is available on GitHub. Some source data is restricted as comes from potentially identifiable sources but aggregate data can be provided by the authors.

https://github.com/terminological/uk-covid-datatools/tree/master/vignettes/serial-interval

## Authors contributions

All authors discussed the concept of the article and RC wrote the initial draft. KTA, EB-P and LD commented and made revisions. All authors read and approved the final manuscript. RC is the guarantor. The views presented here are those of the authors and should not be attributed to TSFT or the GDE.

## Impact of uncertainty in serial interval, generation interval, incubation period and delayed observations in estimating the reproduction number for COVID 19

## Supplementary material

**Supplemental table 2:** Parametererised serial interval distributions from FF100. Gamma and Negative Binomial estimates are from data truncated at zero. AIC estimates are not comparable to those for Normal distribution which is fitting all data, including negative serial intervals, and hence has a lower mean.

| Distribution | AIC | N | Parameter / Moment | Mean ± SD (95% CI) |
| --- | --- | --- | --- | --- |
| gamma | 181 | 43 | shape | $2.74 \pm 0.59$ (1.83; 4.12) |
| | | | rate | $0.77 \pm 0.18$ (0.49; 1.15) |
| | | | mean | $3.56 \pm 0.32$ (3.00; 4.24) |
| | | | sd | $2.18 \pm 0.29$ (1.66; 2.85) |
| negative binomial | 186 | 43 | size | $162.38 \pm 409.00$ (4.08; 1365.73) |
| | | | prob | $0.79 \pm 0.15$ (0.54; 1.00) |
| | | | mean | $3.57 \pm 0.32$ (2.99; 4.07) |
| | | | sd | $2.16 \pm 0.23$ (1.83; 2.59) |
| normal | 251 | 50 | mean | $2.82 \pm 0.40$ (2.12; 3.50) |
| | | | sd | $2.80 \pm 0.30$ (2.25; 3.33) |
| | | | mean | $2.82 \pm 0.40$ (2.12; 3.50) |
| | | | sd | $2.80 \pm 0.30$ (2.25; 3.33) |

**Supplemental figure 1:**
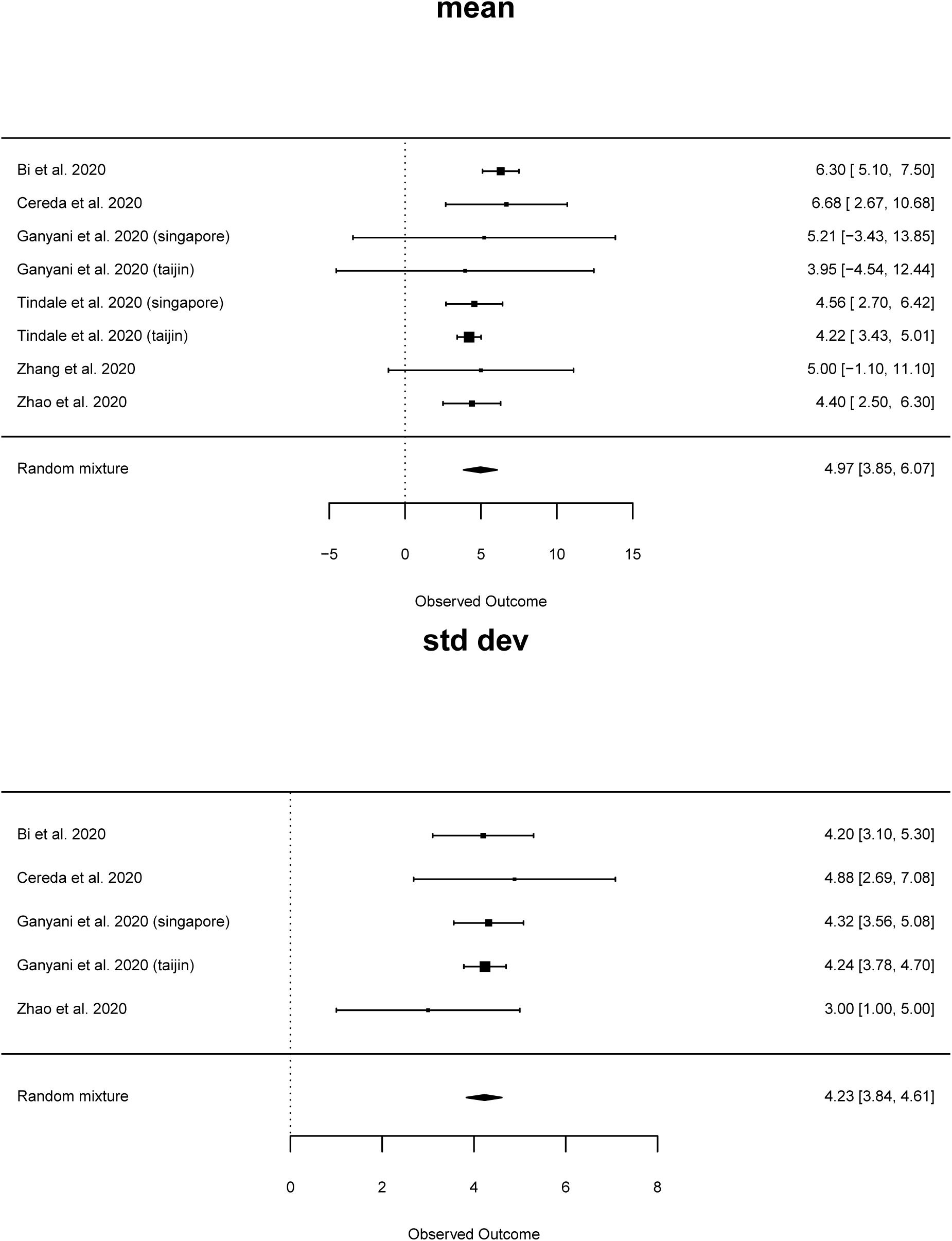
Forest plot for serial interval studies for A, the mean and B, the standard deviation of the serial interval, using the normal mixture random effect model, and from studies identified in the literature which assume a Gamma distributed serial interval.

**Supplemental table 1:** Parametererised serial interval distributions from resampling the literature. Gamma and Negative Binomial estimates are from data truncated at zero. AIC estimates are not comparable to those for Normal distribution which is fitting all data, including negative serial intervals, and hence has a lower mean.

| Distribution | AIC | N | Parameter / Moment | Mean ± SD (95% CI) |
| --- | --- | --- | --- | --- |
| gamma | 6.1e+03 | 113313 | rate | 0.24 ± 0.04 (0.18; 0.30) |
| | | | shape | 1.38 ± 0.24 (1.00; 1.78) |
| | | | mean | 5.61 ± 0.30 (5.13; 6.22) |
| | | | sd | 4.83 ± 0.37 (4.30; 5.69) |
| negative binomial | 6.13e+03 | 113313 | prob | 0.31 ± 0.02 (0.26; 0.35) |
| | | | size | 2.53 ± 0.32 (1.80; 3.10) |
| | | | mean | 5.61 ± 0.30 (5.13; 6.22) |
| | | | sd | 4.26 ± 0.17 (3.98; 4.62) |
| normal | 7.46e+03 | 125439 | mean | 4.83 ± 0.28 (4.31; 5.40) |
| | | | sd | 4.73 ± 0.20 (4.40; 5.15) |
| | | | mean | 4.83 ± 0.28 (4.31; 5.40) |
| | | | sd | 4.73 ± 0.20 (4.40; 5.15) |

**Supplemental table 3:**
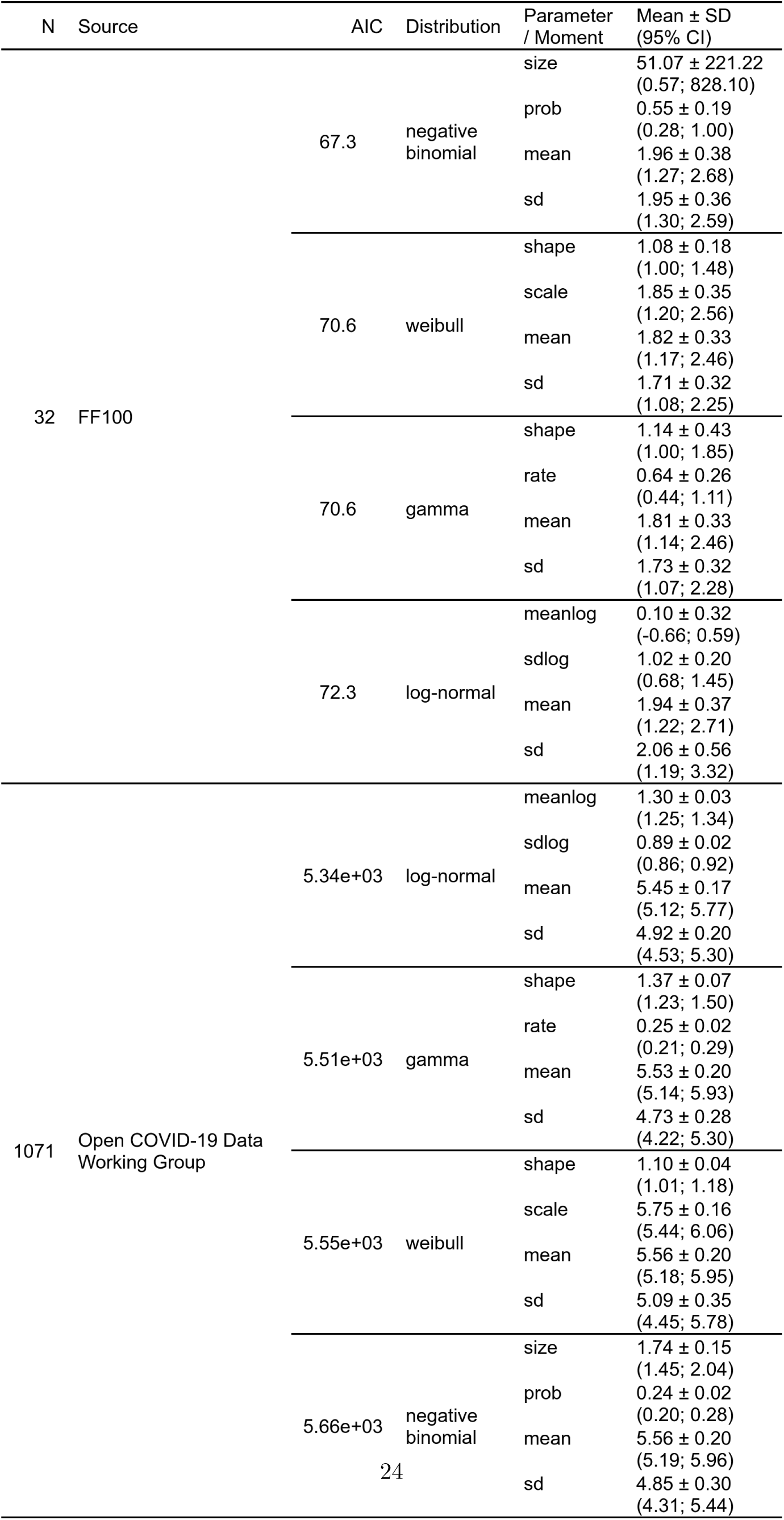
Distribution details for estimated incubation period distributions reconstructed from Open COVID-19 Data Working Group and from FF100 data

**Supplemental table 4:**
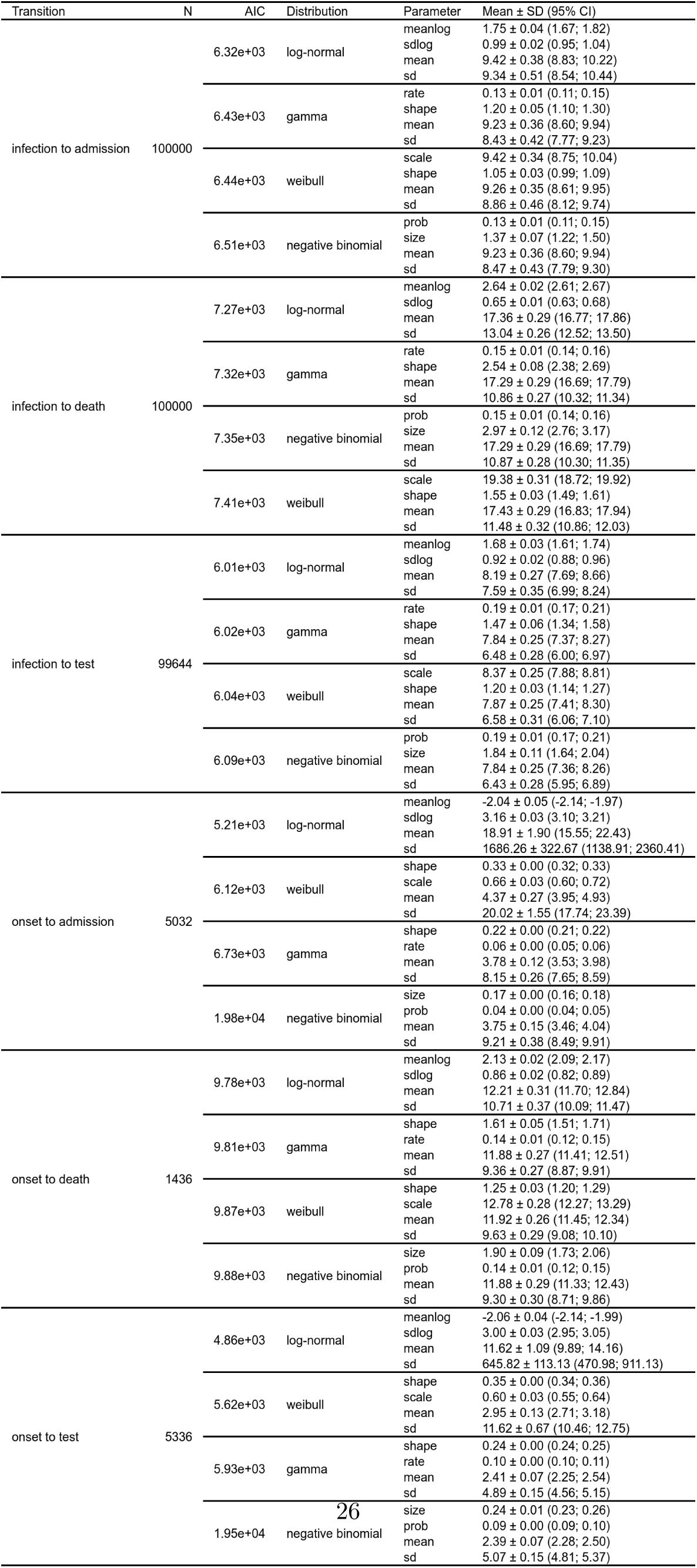
Time delay distributions estimated from CHESS data set, for both transitions from disease onset to case, admission or death, and presumed infection and case, admission or death

**Supplemental figure 2:**
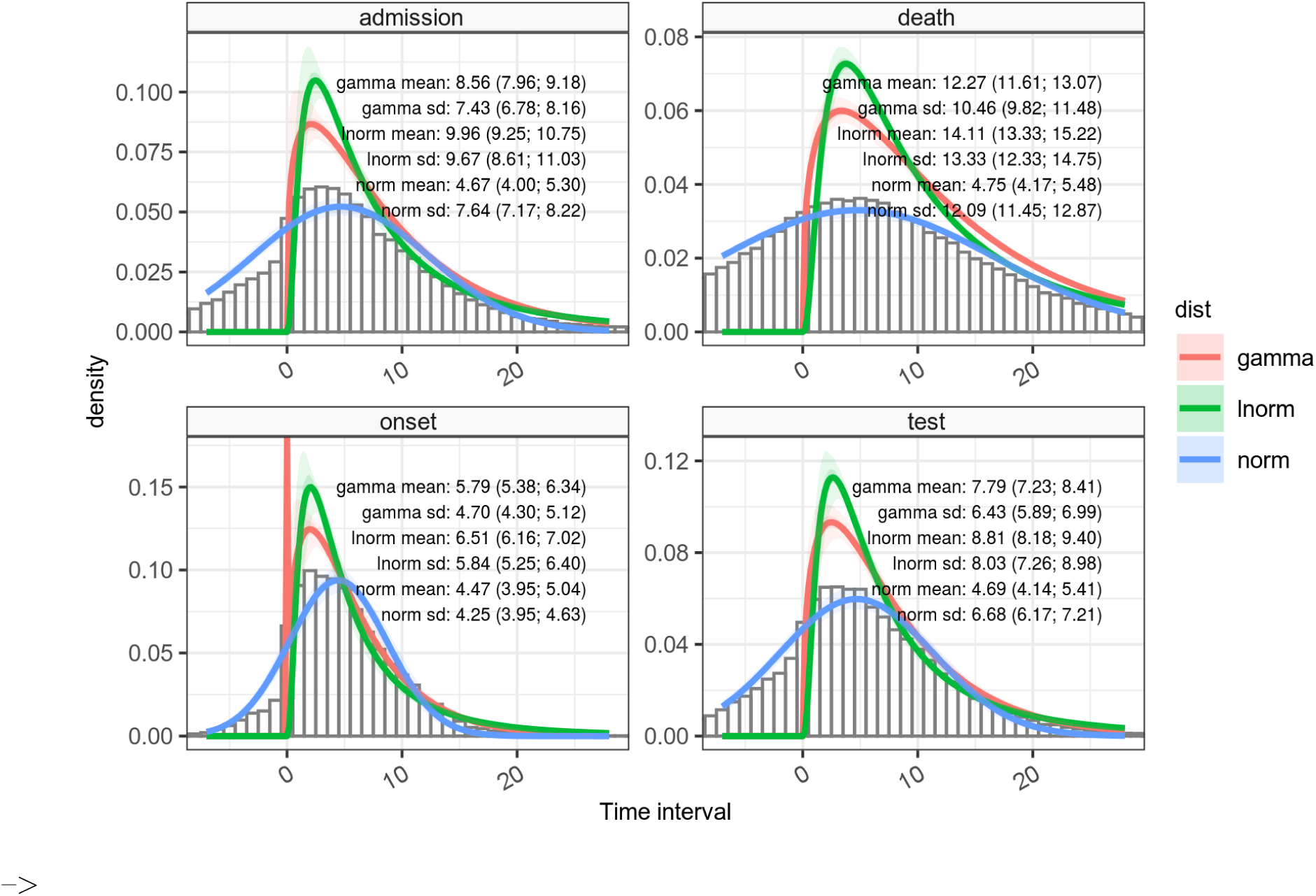
Time intervals between infector-infectee observations.

These distributions are based on the joint probability of serial interval (between onset) estimated from our literature re-sampling analysis and delay distributions from onset to observation, estimated from the CHESS data set. The joint probability is estimated as the combination of bootstrapped samples of raw data assuming these are independent (see discussion section).

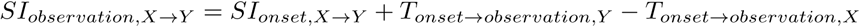

In which the quantity *T*_*onset →observation,Y*_ − *T*_*onset → observation,X*_ is essentially an error term with zero mean. We note that the un-truncated normal distributions mean is approximately equal for all 4 distributions, around 4.5 to 4.8, as the skew of the distribution affects the fitting process. In this figure the distributions were fitted using a maximum goodness of fit estimator. The serial interval for onset estimated in the main paper is included for comparison.

